# The diagnostic accuracy of clinical symptoms and dipstick urinalysis in detecting urinary tract infection among symptomatic patients in Nairobi County, Kenya

**DOI:** 10.1101/2024.05.23.24307793

**Authors:** Hellen Onyango, Derek J Sloan, Katherine Keenan, Mike Kesby, Caroline Ngugi, Humphrey Gitonga, Robert Hammond

## Abstract

**Objectives:** A cross-sectional study was conducted to evaluate the diagnostic performance of clinical symptoms and dipstick urinalysis, both individually and in combination, in predicting urine culture positivity.

**Methods:** Consenting patients with symptoms suggestive of UTI provided midstream urine samples. Dipstick was used to identify Nitrites (NIT) and leucocyte esterase (LE). Urine was cultured on Cystine Lactose Electrolyte Deficient agar (CLED) and incubated at 37^0^C for 24 hours. Pure bacterial colony counts of ≥10^4^ cfu/mL were classified as “positive” for UTI.

**Results:** A total of 552 participants were enrolled. Frequency, smelling cloudy urine, haematuria, and fever were associated with positive urine culture (P <0.05). The symptoms were predictive of UTI when fever was included, and better when a combination of urinary symptoms plus fever were present. Urinary dipstick sensitivity was poor overall. LE+ or NIT+ dipstick result achieved sensitivity of 67%. Combined NIT+ and LE+ had the highest specificity (99%) and a positive predictive value of 91%. Combining symptoms with dipstick improved the predictive accuracy of culture positivity.

**Conclusion:** An algorithm-based combination of symptoms and dipstick testing in outpatient settings where culture is rarely performed could improve UTI diagnosis and promote more appropriate antibiotic use.

## 1. Introduction

UTI is an inflammatory response of the urothelium to bacterial invasion and is among the most frequent community-acquired bacterial infection, affecting more than 150 million individuals globally [1,2]. The disease is associated with morbidity and mortality, particularly among high-risk subpopulations such as children, pregnant women, the elderly and immunocompromised patients [3,4]. Allied to death and disability are prolonged hospital stays, the need for second-line antimicrobial drugs, adverse impacts on the income and livelihoods of individuals, and general economic harm at a national level [5,6].

While prompt diagnosis of UTI is critical for patient management, quantitative urine culture remains the gold standard method for UTI diagnosis despite its widely acknowledged limitations of cost, labour intensity and prolonged time to result of between 24-72 hours [7]. Given these limitaions, clinicians encountering patients UTI-like symptoms often make treatment decisions guided by reported symptoms and the presence of risk factors for clinical complications before the diagnosis is confirmed through urine culture [8]. Some people with UTI present with atypical signs and symptoms, whilst others may have UTI symptoms in the absence of infection, making clinical diagnosis a challenge [9]. Symptoms of uncomplicated UTI include dysuria (pain while passing urine), frequency (frequent voiding of urine), urgency (urge to urinate), haematuria (presence of blood in urine), or passing smelly, cloudy urine (SCU) [10]. While the presence or absence of these typical symptoms may function as a useful UTI screening criterion for UTI, their ability to reliably predict microbial urine culture positivity remains unclear, particularly in resource constrained settings where clinical symptoms are heavily relied upon and culture is rarely performed.

Alongside symptoms, dipstick urinalysis is a rapid point- of - care test routinely used to improve the precision of UTI diagnosis. Dipsticks test markers of infection to a range of medical conditions, based on colorimetric principles. Markers which are mainly helpful to detect UTIs are leucocytes esterase (LE) and nitrite (NIT) which detects bacteriuria or pyuria in urine [11]. Nitrite testing detects the presence of bacteria with enzymes that can convert urinary nitrates to nitrites and is associated with members of the family Enterobacterales. Other urinary pathogens such as *Staphylococcus* spp*., Pseudomonas* spp., and *Enterococcus* spp. do not produce nitrate reductase enzymes [7]. This limits the microbiological range of nitrite-mediated detection of UTI. Another limitation is the fact that a minimum bladder incubation period of 4 hours is required for nitrates to be converted to nitrites at reliably detectable levels [7,9].

The leucocyte esterase (LE) test detects whether leucocytes have produced proteins with esterolytic activity that have then hydrolysed ester substrates. LE testing is likely to give a false positive result when urine is highly contaminated with bacterial vaginal flora; when specimens contain eosinophils or *trichomonas* species, both of which can produce esterase; and when the strip is exposed to an oxidizing agent or formalin [9,12,13]. False negative results may arise when urine has high levels of glycosuria and proteinuria; when urine is preserved using boric acid; and when patient is on antibiotic treatment regimen [12]. In addition to nitrites and leucocyte esterase, other urinary dipstick markers of UTI include blood, increased pH, and proteins. However, these are less specific, as they also have many other causes [8].

A previous meta-analysis conducted on the accuracy of urine dipstick test relative to quantitative urine culture found moderate sensitivity (48%) and specificity (91%) in detecting UTIs. The sensitivity was higher in inpatients (58% vs 45%), while the specificity was greater in outpatients (96% vs 45%)[14]. In High Income Countries (HIC), dipstick urinalysis is widely recognized as a preliminary screening test, providing guidance on whether further culture is necessary. However, in Low-and-Middle Income (LMICs), most hospitals lack the laboratory capacity to perform urine culture and rely entirely on dipstick urinalysis and symptoms for diagnosis [15,16]. This may result in cases of probable misdiagnosis indirectly contributing to delayed or wrong diagnosis, over-treatment or unnecessary treatment of cases with antimicrobials. These factors could in turn lead to the emergence and spread of multidrug resistant clones.

Most previous studies have investigated the accuracy of dipstick urinalysis in detecting UTI without incorporating the patient reported symptoms [15–18]. In contrast, this study systematically evaluates the accuracy of clinical symptoms and dipstick urinalysis, both individually, and in combination when compared to microbiological urine culture in a large outpatient cohort. The study aims to demonstrate how best they can be used despite their limitations, in settings where urine culture is rarely performed.

## 2. Materials and methods

### 2.1. Study design

A cross-sectional study was conducted at Mama Lucy Kibaki Hospital (MLKH) and Mbagathi District Hospital (MDH) between 04/01/2022 to 22/08/2022. The hospitals are located within Nairobi County, Kenya, and mostly serve the urban populace. The Kenyan healthcare system is structured in a hierarchical manner, from community-based primary healthcare services through to specialized hospital care. The current structure consists of six levels (I-VI) in ascending order. Both MLKH and MDH are government owned level (V) tertiary health facilities.

### 2.2. Participant recruitment and sample collection

Adults (≥18 years) and children (5-17 years) were recruited by a resident clinician at the outpatient department of MLKH or MDH if they presented with one or more UTI symptoms (increased urinary frequency or urgency, dysuria, passing strong smelling cloudy urine, haematuria, lower abdominal pain, and/or unexplained fever (≥38°C) in children), or any other feature which made the clinician suspect UTI. Participants were taken through the informed consent document in their preferred language (English or Kiswahili). Written informed consent was obtained from adult patients. Assent and consent were obtained for participants aged 13-17 years. Parents/guardians of participants aged ≤13 years consented on their behalf. Participants/guardians who were unable to sign marked the consent form with a thumb print. Potential participants who did not meet the criteria of a presumptive UTI case, or who declined to consent were excluded from the study. A structured questionnaire was used to collect self-reported socio-demographic details such as age, gender, level of education, monthly income, and prior antibiotic intake. Parents/ guardians of children (5-17 years) filled the questionnaire on their behalf. All data were collected electronically using Epicollect5 mobile application (https://five.epicollect.net) [19]. Consenting participants were instructed on how to aseptically collect clean catch mid-stream urine into a sterile screw capped universal bottle. All samples were assigned a unique study identification number, transported to the microbiology laboratory, and processed within two hours of collection.

### 2.3. Dipstick test

The dipstick test was conducted using combur-10 test M strips according to the manufacturer’s instructions (Combur-10, UK). The urine strip was dipped into approximately 10 ml of the urine specimen, removed immediately, and results read after waiting period of 2 minutes. The strip was held horizontally adjacent to the reagent colour blocks on the strip container and colours carefully matched. LE was reported as negative, trace, 1+ small, 2+ moderate, 3+ large while NIT was recorded as either positive or negative. With reference to the manufacturers guide for interpretation, dipstick testing that produced nitrites or leucocyte esterase greater than trace was taken as positive for UTI. No nitrites or leucocyte esterase were interpreted as negative.

### 2.4. Quantitative urine culture assay

A well-mixed 10 µl urine aliquot was plated directly on CLED (Oxoid, England) and incubated aerobically at 37^0^C for 24 hours. A pure bacterial growth yielding colony counts of 10^4^ cfu/mL was deemed significant for a UTI infection. Mixed urine cultures (with more than one colony type) or those with either low bacterial colony counts of <10^4^ cfu/mL, or without microbial growth were interpreted as negative for UTI. Isolates were identified to the species level using colonial morphological characteristics, Gram-staining technique and standard biochemical tests (catalase, coagulase, urease, oxidase, sulfide indole motility, methyl red, citrate utilization)[20]. Where necessary, Analytical profile index (API 20E) strips, were used to confirm the identity of strains following the manufacture’s guidelines (BioMerieux, Charbonnieres-LesBains, France).

### 2.5. Statistical analysis

Data were downloaded from Epicollect to Microsoft excel for analysis (Microsoft Corp, Red-Mond, Washington, USA). Quantitative data were analysed using STATA Version 16 (StataCorp. College Station, Texas, USA). Categorical variables were presented as counts with proportions, while age was also summarised as median and interquartile range (IQR). Univariable and multivariable logistic regression was performed to assess the relationship between symptoms, and urine culture positivity with the result expressed as an Odds Ratio (OR) and 95% confidence Interval (CI). Categorical variables were analyzed for significant associations using contingency tables and the χ2 test and Fisher exact probability. Statistical significance was considered at probability values less than 0.05.

For urinary dipstick, LE and NIT diagnostic yield were calculated in four ways: LE positive, irrespective of NIT result (LE+), NIT positive irrespective of LE result (NIT+), both NIT and LE positive (NIT+ and LE+), and either NIT or LE positive (NIT+ or LE+) [17]. The sensitivity, specificity, positive predictive value (PPV), negative predictive value (NPV) were calculated with 95 % CI for dipstick parameters using urine culture as the reference standard. Percentage agreement and Cohen’s Kappa coefficient were applied to assess the agreement between dipstick and culture results. [21].

### 2.6. Ethical statement

This study was approved by the University of St. Andrews Teaching and Research Ethics Committee (UTREC), [Approval code. MD15749]; Jomo Kenyatta University of Agriculture and Technology Institutional Ethics Review Board (JKUAT-IERB) [Approval no. JKU/IERC/02316/0166]; National Commission for Science Technology and Innovation (NACOSTI) [Approval no. P/21/12520]. Approvals to access study sites were also obtained from the Nairobi metropolitan services, MLKH and MDH. Each participant in the study provided either a written informed consent or marked the consent form with a thumb print. Although the study involved children, their participation was limited to providing a urine sample under the supervision of their parents/guardians. All questionnaires were completed by parents/guardians on their behalf. The principal investigator was never alone with the children; all interviews were conducted in the presence of an adult. Therefore, certification for working with people in the vulnerable group or the local East African equivalent was not required. All study procedures and experiments were conducted in accordance with the relevant guidelines and regulations.

## 3. Results

### 3.1. Demographic characteristics of study participants

A total of 622 participants were screened for enrolment into the study. However, 552 were enrolled, 402 at Mama Lucy Kibaki Hospital, 150 at Mbagathi District hospital. Seventy were excluded for the reasons outlined in **Fig 1**. The enrolled participants were predominantly young adults with the age group of 21-30 years representing the modal group, and a median age of 29 years (IQR:24-36). There were proportionally more females 72% (398/552). Most of the participants had at least a secondary level of education 270 (49%). Among the 552 individuals recruited, 236 (43%) had taken medication within two weeks preceding their enrolment. Of these individuals, 168 (30%) had been on antibiotics, while the remaining 68 (12%) had taken other types of medication. Details of participants characteristics are shown in **Table 1**.

**Fig 1:**
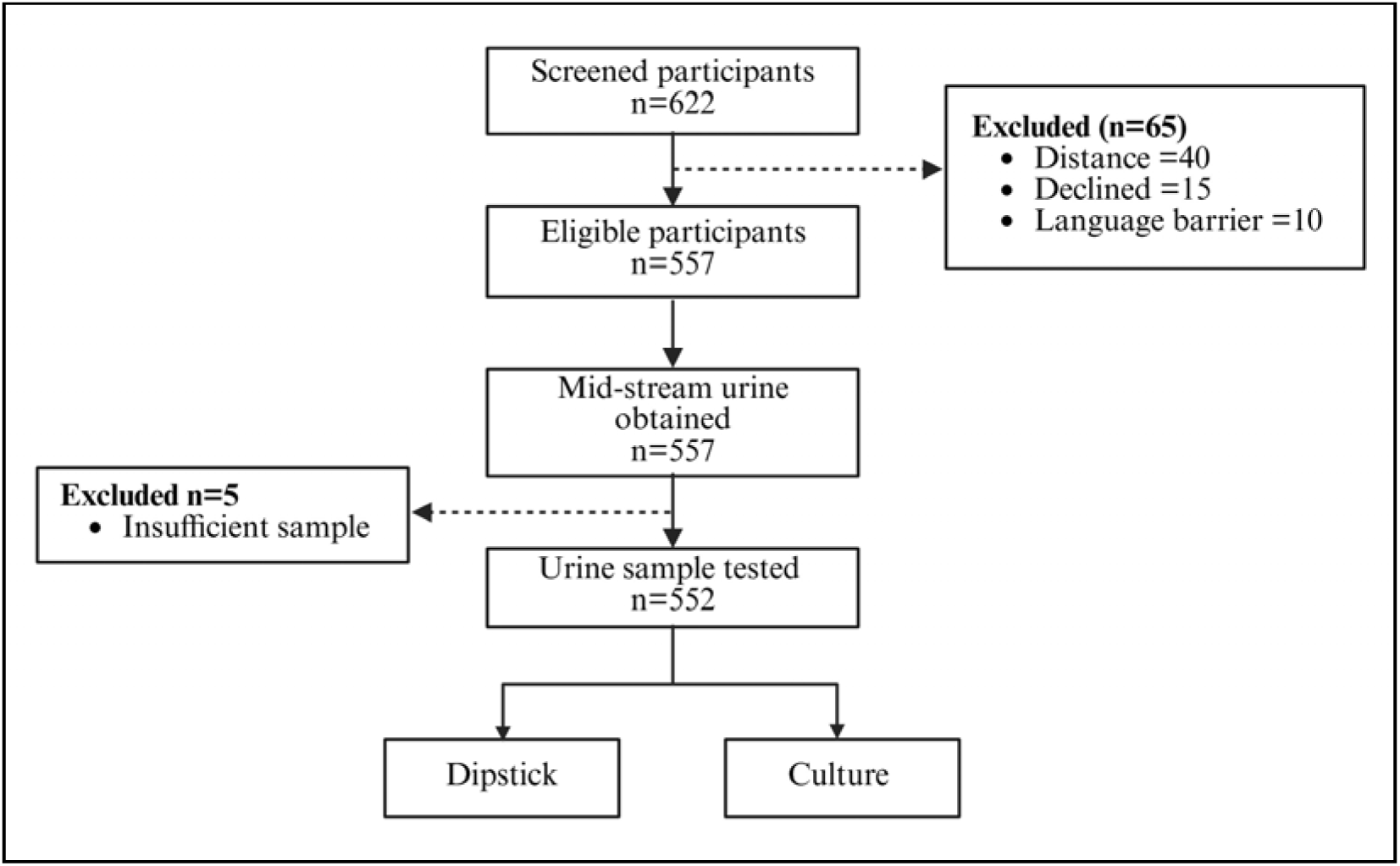
Participant recruitment flow diagram. A total of 622 symptomatic adult and child patients were screened for enrolment. Five hundred and fifty-two urine samples were obtained from eligible participants and tested using dipstick and culture.

**Table 1:**
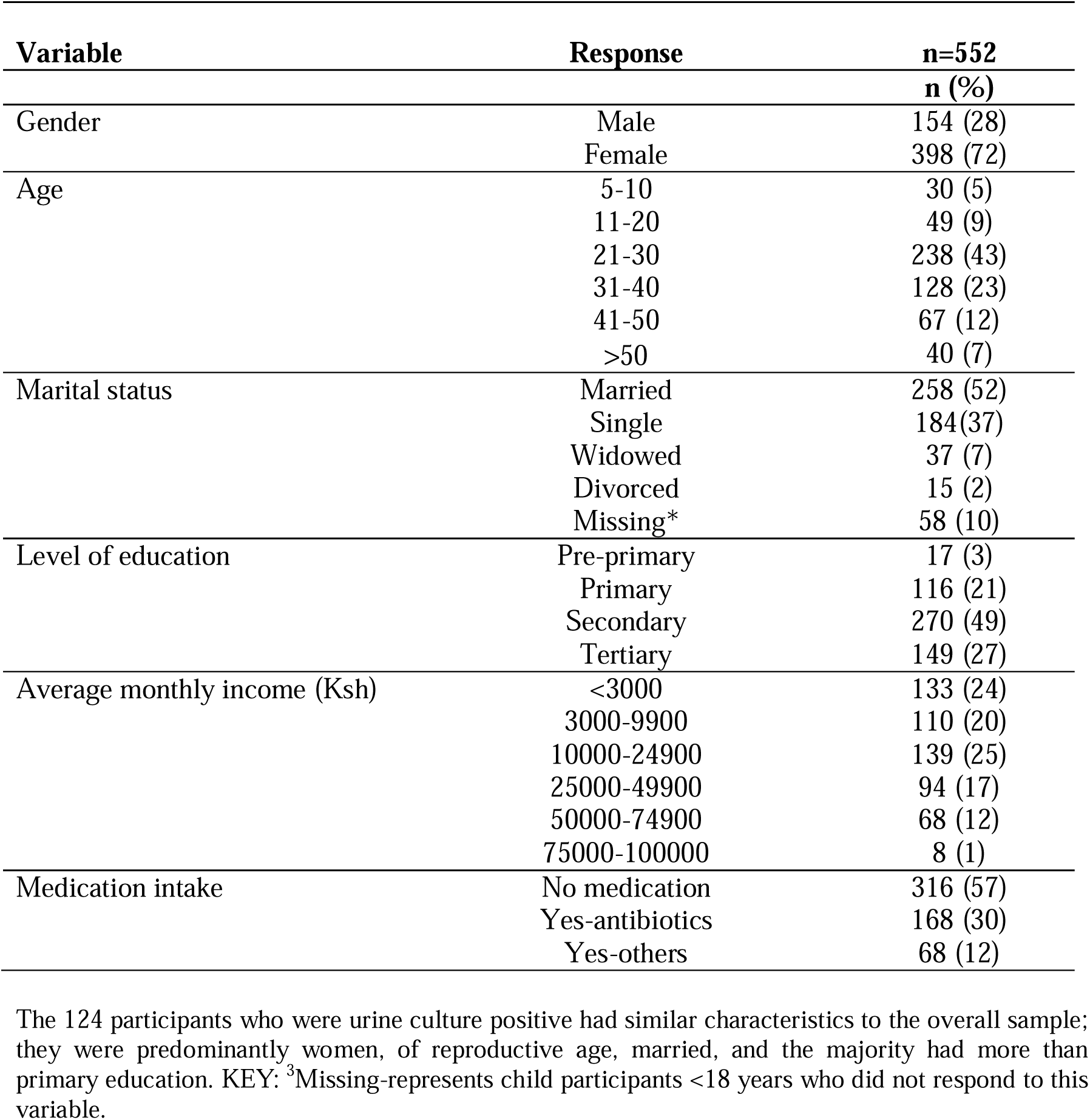
Social-demographic characteristics of study participants.

### 3.2. Microbiological characteristics

Among 552 urine samples analysed, 124 (22%) were positive for UTI (a monoculture bacterial growth of ≥10^4^ cfu/mL). The proportion of infections attributable to Gram-negative organisms were 97 (78%). The most prevalent uropathogen was *Escherichia coli*, 64 (51.6%). *E. coli* was followed in order of prevalence by *Klebsiella pneumoniae*, 16 (12.9%); *Staphylococcus aureus*, 14 (11.3%); Coagulase Negative Staphylococcus (CoNS), 7 (5.6%); *Enterococcus faecalis*, 6 (4.8%), *Klebsiella* spp., 5 (4%); *Proteus mirabilis,* 5 (4%); and *Acinetobacter baumanni*, 1(0.8%). *Pseudomonas aeruginosa*, *Proteus vulgaris*, and *Citrobacter koseri* each were 2 (1.6%).

### 3.3 Accuracy of clinical symptoms in detecting UTI

The clinical symptoms were grouped into specific urinary symptoms and systemic/other symptoms. Specific urinary symptoms included dysuria, frequency, urgency, passing smelling cloudy urine (SCU), and haematuria. Systemic/other symptoms included fever (>38°C), lower abdominal pain or tenderness (LAP/T), urethral/vaginal discharge (U/VD), vomiting, backpain, painful sexual intercourse (PSI), and miscellaneous others (e.g., blisters on penis/pubic area, irritability, unspecified). More than half of the participants 292 (53%) presented with urinary symptoms only, of which 56 (19%) were confirmed urine culture positive, 85 (15%) had systemic/other symptoms only, whilst 175 (32%) had both urinary and systemic/other symptoms.

Based on univariable logistic regression analysis of individual symptoms **Table 2**, frequency (OR 2.1, 95% CI: 1.20-3.64), SCU (OR 3, 95% CI: 1.76-5.36), haematuria (OR 3, 95% CI: 1.01-9.27), fever (OR 3, 95% CI; 1.99-4.76) were associated with a microbiological confirmation of UTI on urine culture. A set of multivariable model was fitted to determine which individual symptom or combination of symptoms could predict positive urine culture results most effectively. All the symptoms were included irrespective of the univariable analysis outcome in **Table 2**. The symptoms were grouped into urinary symptoms, fever as a general marker of infection, and other reported symptoms. From the 3 main groups, 7 combinatory categories were generated **Fig. S1,** and used as predictors in a multivariable model adjusted for age and gender, to assess how they were associated with positive urine culture results, **Table 3**. Predictive margins were calculated to determine the probability of the % individuals positive for urine culture when these combined symptoms were present.

**Table 2:** Clinical characteristics of study participants (n=552)

| Characteristics | Culture results |  |  |  |  |  |
| --- | --- | --- | --- | --- | --- | --- |
|  | Frequency<br>n (%) | Negative<br>n=428 | Positive<br>n=124 | OR | 95% CI | p-value |
| Specific urinary symptoms |  |  |  |  |  |  |
| Dysuria |  |  |  | 1.41 | 0.91-2.05 | 0.12 |
| No | 274 (50) | 220 | 54 |  |  |  |
| Yes | 278 (50) | 208 | 70 |  |  |  |
| Urgency |  |  |  | 0.78 | 0.48-1.26 | 0.32 |
| No | 413 (75) | 316 | 97 |  |  |  |
| Yes | 139 (25) | 112 | 27 |  |  |  |
| Frequency |  |  |  | 2.12 | 1.20-3.64 | 0.01 |
| No | 487 (88) | 386 | 101 |  |  |  |
| Yes | 65 (12) | 42 | 23 |  |  |  |
| SCU |  |  |  | 3.07 | 1.76-5.36 | <0.001 |
| No | 492 (89) | 394 | 98 |  |  |  |
| Yes | 60 (11) | 34 | 26 |  |  |  |
| Haematuria |  |  |  | 3.08 | 1.01-9.27 | 0.048 |
| No | 539 (98) | 421 | 118 |  |  |  |
| Yes | 13 (2.5) | 7 | 6 |  |  |  |
| Systemic/other symptoms |  |  |  |  |  |  |
| LAP/T |  |  |  | 1.07 | 0.68-1.69 | 0.76 |
| No | 411 (74) | 320 | 91 |  |  |  |
| Yes | 141 (26) | 108 | 33 |  |  |  |

**Table 2: Continued**
| **Characteristics** | **Culture results** |  |  | **OR** | **95% CI** | **p-value** |
| --- | --- | --- | --- | --- | --- | --- |
|  | **Frequency**<br>**n (%)** | **Negative**<br>**n=428** | **Positive**<br>**n=124** |  |  |  |
| **Fever** |  |  |  | 3.08 | 1.99-4.76 | <0.001 |
| No | 425 (77) | 351 | 74 |  |  |  |
| Yes | 127 (23) | 77 | 50 |  |  |  |
| **U/VD** |  |  |  | 0.68 | 0.23-1.65 | 0.34 |
| No | 520 (94) | 401 | 119 |  |  |  |
| Yes | 32 (6) | 27 | 5 |  |  |  |
| **Vomiting** |  |  |  | 8.2 | 2.80-24.1 | <0.001 |
| No | 536 (97) | 423 | 113 |  |  |  |
| Yes | 16 (3) | 5 | 11 |  |  |  |
| **Back pain** |  |  |  |  |  | 0.98 |
| No | 542 (98) | 418 | 124 | - | - |  |
| Yes | 10 (2) | 10 | 0 |  |  |  |
| **Painful sexual intercourse** |  |  |  |  |  | 0.98 |
| No | 542 (98) | 418 | 124 | - | - |  |
| Yes | 10 (2) | 10 | 0 |  |  |  |
| **Others** |  |  |  | 4.7 | 1.04-21.3 | 0.04 |
| No | 545 (98.7) | 425 | 120 |  |  |  |
| Yes | 7 (1.3) | 3 | 4 |  |  |  |

Among the symptoms, Frequency, SCU, haematuria, and fever were significantly associated with positive urine culture results ( $P < 0.05$ ). KEY: LAP/T-Lower abdominal pain or tenderness, SCU-Strong smelling cloudy urine, U/VD-Urethral/ vaginal discharge, Others included blisters on penis/pubis area 2 (0.3%), irritability 1 (0.2%), unspecified 4 (0.7%).

**Table 3:**
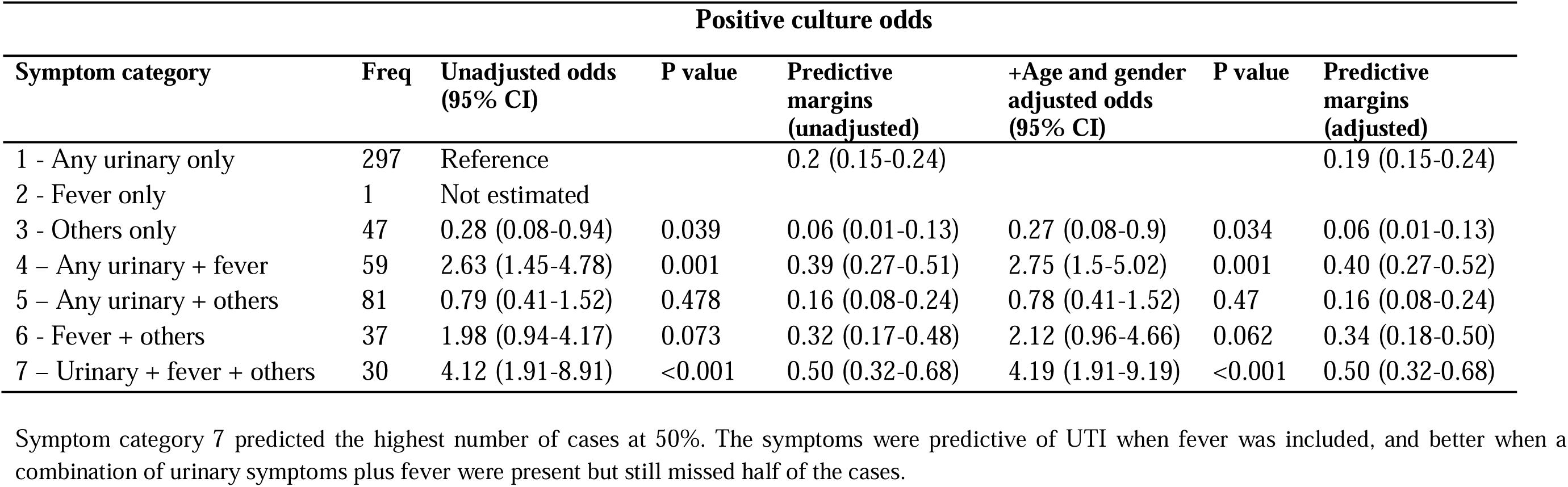
Association between symptom categories and urine culture results (n=552)

| Positive culture odds |  |  |  |  |  |  |  |
| --- | --- | --- | --- | --- | --- | --- | --- |
| Symptom category | Freq | Unadjusted odds (95% CI) | P value | Predictive margins (unadjusted) | +Age and gender adjusted odds (95% CI) | P value | Predictive margins (adjusted) |
| 1 - Any urinary only | 297 | Reference |  | 0.2 (0.15-0.24) |  |  | 0.19 (0.15-0.24) |
| 2 - Fever only | 1 | Not estimated |  |  |  |  |  |
| 3 - Others only | 47 | 0.28 (0.08-0.94) | 0.039 | 0.06 (0.01-0.13) | 0.27 (0.08-0.9) | 0.034 | 0.06 (0.01-0.13) |
| 4 – Any urinary + fever | 59 | 2.63 (1.45-4.78) | 0.001 | 0.39 (0.27-0.51) | 2.75 (1.5-5.02) | 0.001 | 0.40 (0.27-0.52) |
| 5 – Any urinary + others | 81 | 0.79 (0.41-1.52) | 0.478 | 0.16 (0.08-0.24) | 0.78 (0.41-1.52) | 0.47 | 0.16 (0.08-0.24) |
| 6 - Fever + others | 37 | 1.98 (0.94-4.17) | 0.073 | 0.32 (0.17-0.48) | 2.12 (0.96-4.66) | 0.062 | 0.34 (0.18-0.50) |
| 7 – Urinary + fever + others | 30 | 4.12 (1.91-8.91) | <0.001 | 0.50 (0.32-0.68) | 4.19 (1.91-9.19) | <0.001 | 0.50 (0.32-0.68) |

Symptom category 7 predicted the highest number of cases at 50%. The symptoms were predictive of UTI when fever was included, and better when a combination of urinary symptoms plus fever were present but still missed half of the cases.

The best performing symptom categories were category 7 (urinary + fever + others) which predicted positive UTI at 50% of cases, and category 4 (urinary + fever) which predicted positive results 39% of the time. Category 3 (other symptoms only) predicted positive UTI in just 6% of cases, while category 1 (urinary only) had a predictive probability of 20%. Age and gender did not change the associations between symptoms and urine culture, suggesting they were not strong confounders of the relationship, **Table 3**.

### 3.4 Diagnostic accuracy of dipstick using urine culture as a reference

The dipstick positivity rate was dependent on how LE+ and NIT+ results were interpreted. If ‘NIT+ or LE+’ was considered, 187 (34%) of participants were positive, translating to a higher positivity rate than culture. If only ‘LE+’ was used, 158 (29%) were positive which also exceeded the culture positivity rate. ‘NIT+’ and ‘NIT and LE+’ analyses yielded 63 (11%) and 34 (6%) positive results respectively which were lower than the culture positivity rate. In general, dipstick positivity was higher when based on ‘LE+’ alone, and lower when a ‘NIT+’ result was required.

Overall, the diagnostic sensitivity (95% CI) of the urine dipstick test in relation to the gold standard of urine culture, was poor ranging from 25%-67%, **Table 4**. Having either NIT+ or LE+ yielded the highest sensitivity (95% CI) of 67% (57.8 – 74.9), and NPV of 88.8% (84.9–91.7), but a low PPV of 44.4 (37.2 – 51.8). The combination of NIT+ and LE+ results yielded the lowest sensitivity (95% CI) of 25 % (17.8 – 33.7) but the highest specificity (95% CI) of 99% (97.8 – 99.8) and PPV of 91% (75.2 – 97.7). NIT+ alone had a low sensitivity (95% CI) of 43.5% (34.8 – 52.7) and a specificity (95% CI) of 97.9% (95.9 – 98.8). The specificity of urine dipstick was higher when NIT+ was required as part of a positive result. None of age, gender or use of prior medication, including antibiotics, substantially altered any performance characteristics of urinary dipstick, with the exception of LE+ where sensitivity and PPV varied by age and prior antibiotic use, **Tables S1-S4.**

**Table 4:** Diagnostic performance of NIT and LE relative to quantitative urine culture.

| **Dipstick parameter** | **Performance characteristics (95% CI)** |  |  |  |  |  |  |  |
| --- | --- | --- | --- | --- | --- | --- | --- | --- |
|  | **TP** | **FP** | **TN** | **FN** | **Sensitivity %**<br>**(95% CI)** | **Specificity %**<br>**(95% CI)** | **PPV %**<br>**(95% CI)** | **NPV %**<br>**(95% CI)** |
| NIT+ | 54 | 9 | 419 | 70 | 43.5 (34.8 – 52.7) | 97.9 (95.9 – 98.8) | 85.7 (74.1 – 92.9) | 85.7 (82.2 – 88.6) |
| LE+ | 60 | 98 | 330 | 64 | 48.4 (39.4 – 57.5) | 77.1 (72.8 – 80.9) | 37.9 (30.5 – 46.1) | 83.8 (79.7 – 87.2) |
| NIT+ and LE+ | 31 | 3 | 425 | 93 | 25.0 (17.9 – 33.7) | 99.3 (97.8 – 99.8) | 91.2 (75.2 – 97.7) | 82.1 (78.4 – 85.2) |
| NIT+ or LE+ | 83 | 104 | 324 | 41 | 66.9 (57.8 – 74.9) | 75.7 (71.3 – 79.6) | 44.4 (37.2 – 51.8) | 88.8 (84.9 – 91.7) |
| Dipstick performance was generally poor. Using either LE+ or NIT+ achieved the highest sensitivity of 67%. Combined NIT+ and LE+ had the highest specificity and a positive predictive value of 91%. NIT: Nitrites, LE: Leucocyte esterase, TP: True positive, TN: True negative, FP: False positive, FN: False negative, PPV: Positive predictive value, NPV: Negative predictive value. |  |  |  |  |  |  |  |  |

### 3.5 Accuracy of symptoms in combination with dipstick

Since clinicians often use symptoms in combination with dipstick results when making decisions, the performance of the 7 symptom categories outlined in **Fig**. **S1** were evaluated in combination with a dipstick result of either NIT+ or LE+ (chosen because it had the highest sensitivity rate). Similar to results displayed in **Table 3**, Category 7 (all symptoms), and category 4 (urinary + fever), when combined with dipstick were associated with the highest odds of positive urine culture results, and the highest predictive margins, both over 80%, **Table 5**. Comparing the predictive values in **Table 3** and **5**, adding a dipstick result enhanced the predictive probability, compared with using symptoms alone, but did not change which symptoms were more or less predictive.

**Table 5:** Combination of symptoms and dipstick urinalysis in UTI detection.

| **Symptom + dipstick (NIT+ or LE+)** |  | **Positive culture odds** |  |  | **Predictive margins** |  |
| --- | --- | --- | --- | --- | --- | --- |
| **Symptoms** | **Frequency** | **OR** | **95% CI** | **P-value** | **Margin** | **95% CI** |
| 1 - Any urinary only | 297 | Reference |  |  | 0.36 | 0.26-0.45 |
| 2 - Fever only | 1 | - | - | - | - | - |
| 3 - Others only | 47 | 1.4 | 0.37-5.85 | 0.56 | 0.30 | 0.11-0.58 |
| 4 -Any urinary + fever | 59 | 16.8 | 5.55-51.1 | 0.01 | 0.81 | 0.64-0.97 |
| 5 -Any urinary + others | 81 | 1.8 | 0.87-3.71 | 0.11 | 0.33 | 0.17-0.48 |
| 6 - Fever + others | 37 | 5.8 | 1.87-18.2 | 0.02 | 0.61 | 0.35-0.87 |
| 7 -Any urinary + fever + others | 30 | 29.4 | 1.90-8.90 | 0.01 | 0.88 | 0.68-1.09 |

Category 7 and category 4 combined with dipstick were the best predictors of UTI with margins above 80%. In contrast to when used alone, when combined with dipstick, the predictive margin of any urinary symptoms only improved to 36% from 20%.

## 4.0 Discussion

In many resource constrained settings, patient symptoms in combination with dipstick urinalysis are routinely used as a basis for UTI diagnosis and antibiotic prescriptions. This study evaluated the accuracy of patient reported symptoms and dipstick urinalysis, both individually, and in combination, in predicting a positive urine culture, particularly to inform their use in settings where confirmatory culture may not be available. The overall results show that symptoms are predictive of UTI when fever is included, and better when combinations of urinary symptoms plus fever were present but still misses half of confirmed cultures cases. Dipstick urinalysis was poor at predicting positive urine cultures, but combining dipstick with symptoms increased the probability of case detection.

Even though all 552 participants recruited to this study had typical symptoms of UTI, only 124 (23%) were confirmed positive by urine culture, illustrating the difficulty for healthcare providers in deciding on the need for antibiotic prescription based on clinical presentation alone. In this study, individual symptoms such as frequency, SCU, haematuria and fever showed modest diagnostic value. The finding of SCU as a predictor of positive urine culture (P<0.001) is consistent with previous studies which reported urine cloudiness/turbidity as good predictor of UTI.[22,23] Visual evaluation of urine cloudiness/ turbidity has been recommended in some guidelines as part of a process in the diagnosis of uncomplicated UTIs.[24,25] Application of this criterion in settings where microbiology diagnostics are limited, may assist responsible antibiotic use.

While dysuria was the most common individual symptom reported, it was not significantly associated with microbiological confirmation of UTI. In a systematic review of 16 studies incorporating 3,711 patients, on the diagnostic accuracy of UTI symptoms, Giesen *et al*., reported that no symptom on its own has power to rule in or out UTI.[26] However, the predictive value of a symptom may vary depending on how well the patient describes the symptom, the examining physician and the definition of the clinical terminology.[26] This study has shown that combination of multiple symptoms such as any urinary + fever + other symptoms are more predictive of positive urine culture than individual symptoms in isolation. The value of combined symptom is useful, however, no matter how many symptoms are considered, their predictive margins don’t go beyond 50%, and so whilst helpful, this approach is not enough by itself. Consideration of larger and more diverse samples would be needed to confirm these results, but the development of a clinical symptom algorithm may aid clinicians in settings where urine culture remains unavailable as a diagnostic test.

The urine dipstick test is a complex read-out with different variables reported. In this study, interpreting dipstick based on NIT+ test alone had a high specificity but low sensitivity. In prior studies, the presence of nitrites in urine has been shown to be highly specific for bacteriuria (96.6%-97.5%), but has a low sensitivity of 0-44% for bacteriuria between 10^3^-10^5^ cfu/ml [9,11,17,27,28]. The typically low sensitivity value for nitrites can be explained by the process of nitrification that requires approximately 4 hours for detectable nitrite levels to be produced in urine [29]. Alternative explanations could be the lack of dietary nitrates, dilution of the nitrite in urine with diuretics, and unreliability of the nitrite test in detecting Gram-positive organisms [7]. Therefore, the presence of nitrites is highly predictive of UTI, but their absence does not exclude it.

The LE+ test alone has a slightly higher sensitivity, and lower specificity when compared to the individual performance of NIT+. The LE sensitivity of 62.2% reported by Maina *et al* [16], 60% by Dadzie *et al* [17] and 48% by Anith *et al* [11] were all higher than the nitrite test, which mirrors that of the present study. Despite being more sensitive than nitrites, the LE test is not specific for UTI and may be associated with other inflammatory disorders affecting the urinary tract such as vaginitis, chlamydial urethritis or other infections than can elicit an immune response and production of white blood cells [30]. These factors may have contributed to variability in the positive predictive value of LE test from 19% to 88% in prior work, and may also undermine its usefulness in the present dataset [27,31].

In the context of clinical decision-making for infection management, diagnostic tests can be used to ‘rule-out’ or to ‘rule-in’ the need for antibiotic therapy. From these data, the requirement for both NIT+ and LE+ to confer dipstick positivity had potential as a good ‘rule-out’ test due to its high specificity of 99%, albeit with a cautionary note to clinicians that of 518 patients who were dipstick negative on this basis, 93 (17.9%) had positive urine cultures. Most uncomplicated UTIs are self-limiting in large patient groups [32], suggesting that an initial missed diagnosis will often have minimal consequences. However, for certain patient groups the balance of risk may be different; e.g., for pregnant women, a missed UTI diagnosis could lead to poor perinatal and maternal outcomes [33]. Overall, the findings concur with a meta-analysis that support the use of urine dipstick test in a rule-out strategy[34]. A similar conclusion was drawn by Deville *et al* in a meta-analysis that included 70 publications, despite a considerable variation in settings of the different studies that included both out- and in-patients, emergency department, ante-natal unit, and all levels of care from the community to tertiary care [14]. These results are all contradictory to an older meta-analysis which reported that in many clinical settings, the probability of a positive culture with a negative dipstick test is too high to dismiss the probability of a UTI [35].

An alternative, and common, use of the urine dipstick test in resource limited settings is by interpreting the positive results as a tool to support the diagnosis of UTI (rule-in strategy) and as the key result necessary to justify to antibiotic initiation. Whilst ‘either NIT+ or LE+’ confers the highest sensitivity for any dipstick positivity analysis in this study, the result of 67% sensitivity is still unsatisfactory (1/3 of patients with culture confirmed UTI will be missed), and the specificity was low. Additionally, the specificity of interpreting the dipstick result in this was low. These findings mirror those of Maina *et al* and Dadzie *et al* who report an overestimation of UTI by ‘either NIT+ or LE+’ test results compared to culture by 74% and 72% respectively. The consequence may be that some patients who don’t need antibiotics will receive them, whilst a concerning proportion of those who need them may not have them prescribed. On the other hand, this study findings indicate that combining dipstick markers (NIT+ or LE+) with clinical symptoms improved the prediction of positive urine culture results, which accords with prior literature.[8,36] for a combination of all the category of symptoms, or urinary + fever, the predictive margins were ≥80, thus, in settings with poor availability of microbiology diagnostics, these methods appear reasonable in combination for use in UTI diagnosis. The overall context highlights the need for development and expanded availability of new, reliable point of care tests to guide decision-making in UTI management. This study has some limitations. The study was conducted in two purposively chosen health facilities in an outpatient setting. Generalisation to other settings or patient populations can be challenging, particularly when symptoms associated with UTI may stem from other underlying medical conditions. Despite the enrolment of over 500 participants, the number of positive reference test based on culture was limited by antibiotic intake prior to recruitment into the study and a definition of positivity that required a concentration of 10^4^cfu/mL. The findings suggesting that combining dipstick urinalysis with clinical symptoms under particular algorithms would aid clinicians in diagnosing UTI should be investigated with larger samples and sub-group of patients.

## 5.0. Conclusion

The data presented herein demonstrate the need to improve upon the current rapid methods for UTI diagnosis. While symptom patterns are important in giving an indication of how likely UTI confirmation will be, using them in isolation leads to missing half of the true cases. The overall results show that urine dipstick test is an inadequate tool to assess the probability of a positive urine culture. However, given the test characteristics, the dipstick test may be considered as a rule-out tool for antibiotic prescriptions; a negative dipstick test result characterized by no reaction for both nitrites and leucocyte esterase has very high specificity to predict a negative urine culture, making the diagnosis of UTI unlikely and empirical antimicrobial therapy unnecessary. Evaluation of symptoms alongside dipstick results improve UTI detection rates yet remains insufficient because the overall diagnostic performance is sub-optimal. The challenges of performing urine culture diagnostics in resource limited settings, calls for research to develop and validate novel near or point of care technologies with better performance to assist clinicians in clinical decision making. The study provides robust evidence in a real-world LMIC outpatient setting that can change how UTI screening is performed.

## Data Availability

All data produced in the present study are available upon reasonable request to the corresponding author

## Acknowledgements

We sincerely thank all the participants for their valuable participation and cooperation, which made this research possible. We are also grateful to MLKH and MDH for granting us the opportunity to conduct this research, not forgetting the clinicians and laboratory personnel whose support was invaluable during data collection.

## Author contributions

HO, RH, DS, MK, KK.: Conceptualized the study. HO.: coordinated participant recruitment, performed laboratory investigations, data curation, formal data analysis and interpretation, and original drafting of the manuscript. RH, DS, MK, KK.: supervised, reviewed and edited manuscript. DS. Provided clinical expertise on results interpretation. KK. Supported data analysis and interpretation. CN.: Supervised data collection in Kenya. HG.: Supported laboratory investigations and data curation. All authors read and approved submission of the manuscript.

## Funding

This work was supported by the Scottish Funding Council through the Global Challenges Research Fund (GCRF).

## Competing interests

The authors declare no competing interests.

## Data availability

All datasets generated or analysed during this study are available on the University of St Andrews One drive and is accessible upon reasonable request from the corresponding author and meeting the ethical requirements as per consent received from study participants. Please contact for data access.

## SUPPLEMENTARY MATERIALS

**Fig. S1:**
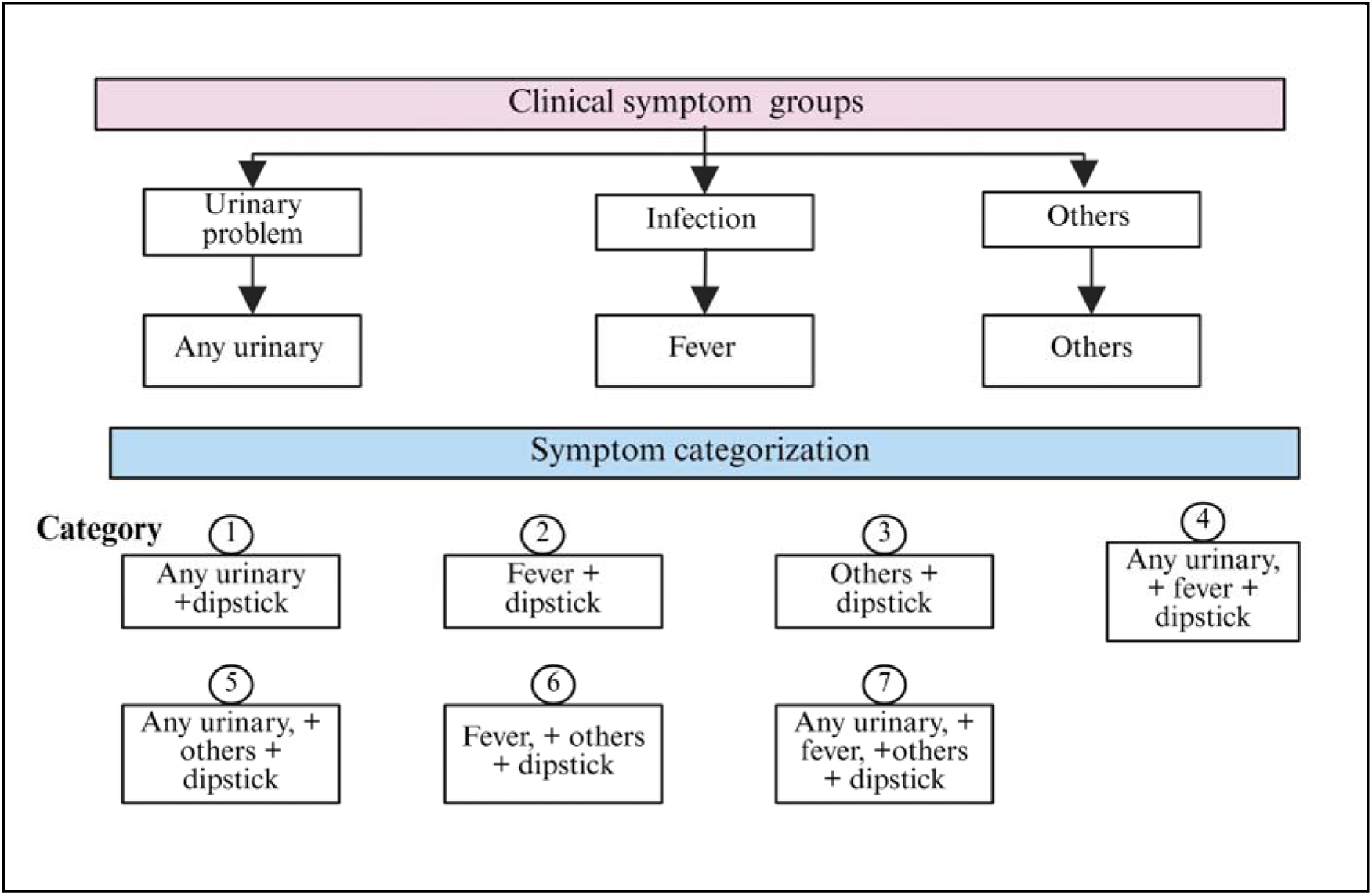
Symptom categories for multivariable regression analysis. The symptoms were grouped into three groups: urinary symptoms (dysuria, frequency, urgency, haematuria, smelly cloudy urine), fever was used as a marker of infection, and other symptoms (Lower abdominal pain or tenderness, U/VD-Urethral/vaginal discharge, vomiting, back pain, painful sexual intercourse, blisters on penis/pubic area, irritability, unspecified). From the 3 groups, 7 combinatory categories were generated as shown and used as predictors in a multivariable regression model.

**Tables S1-S4:**
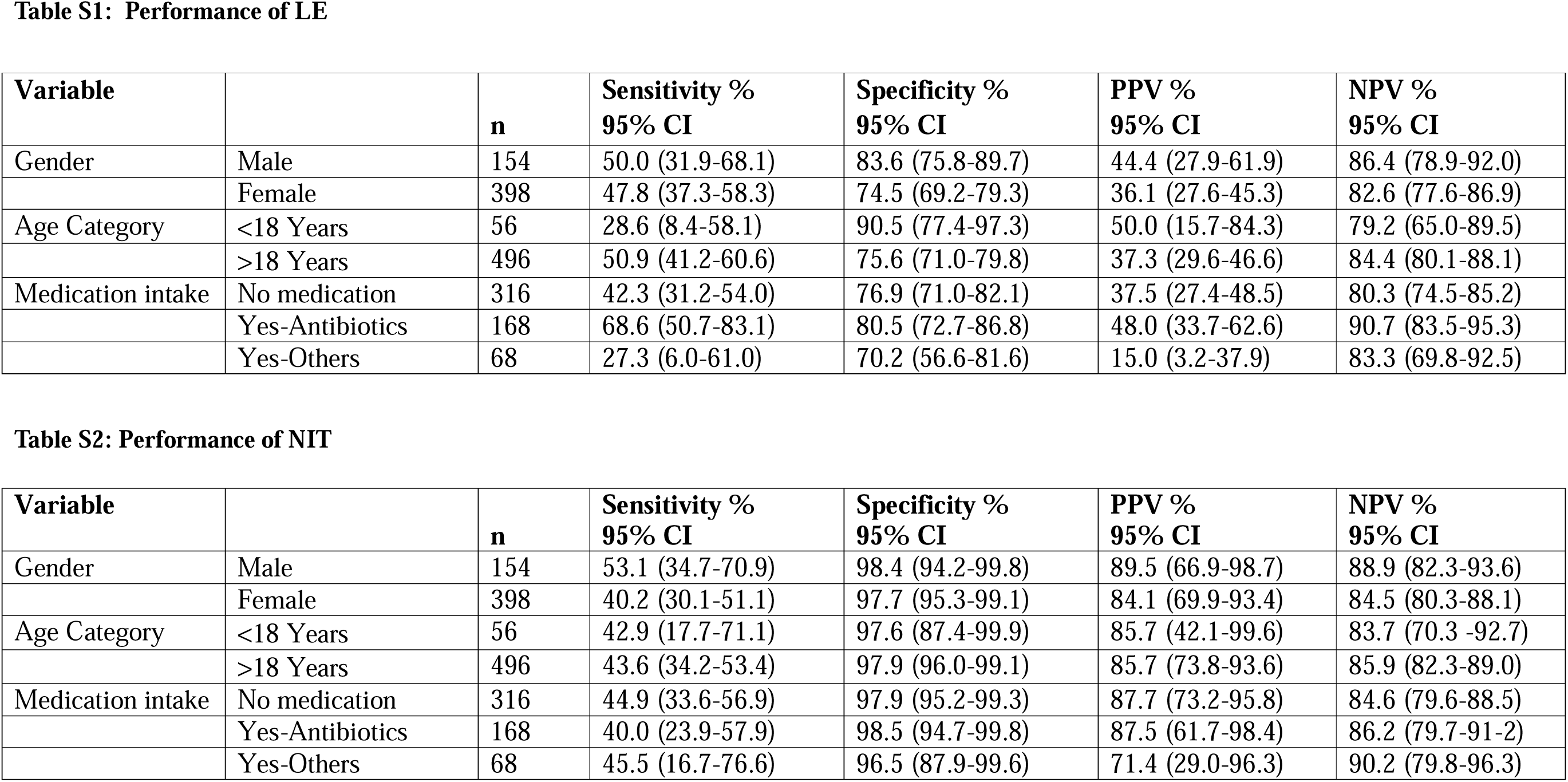
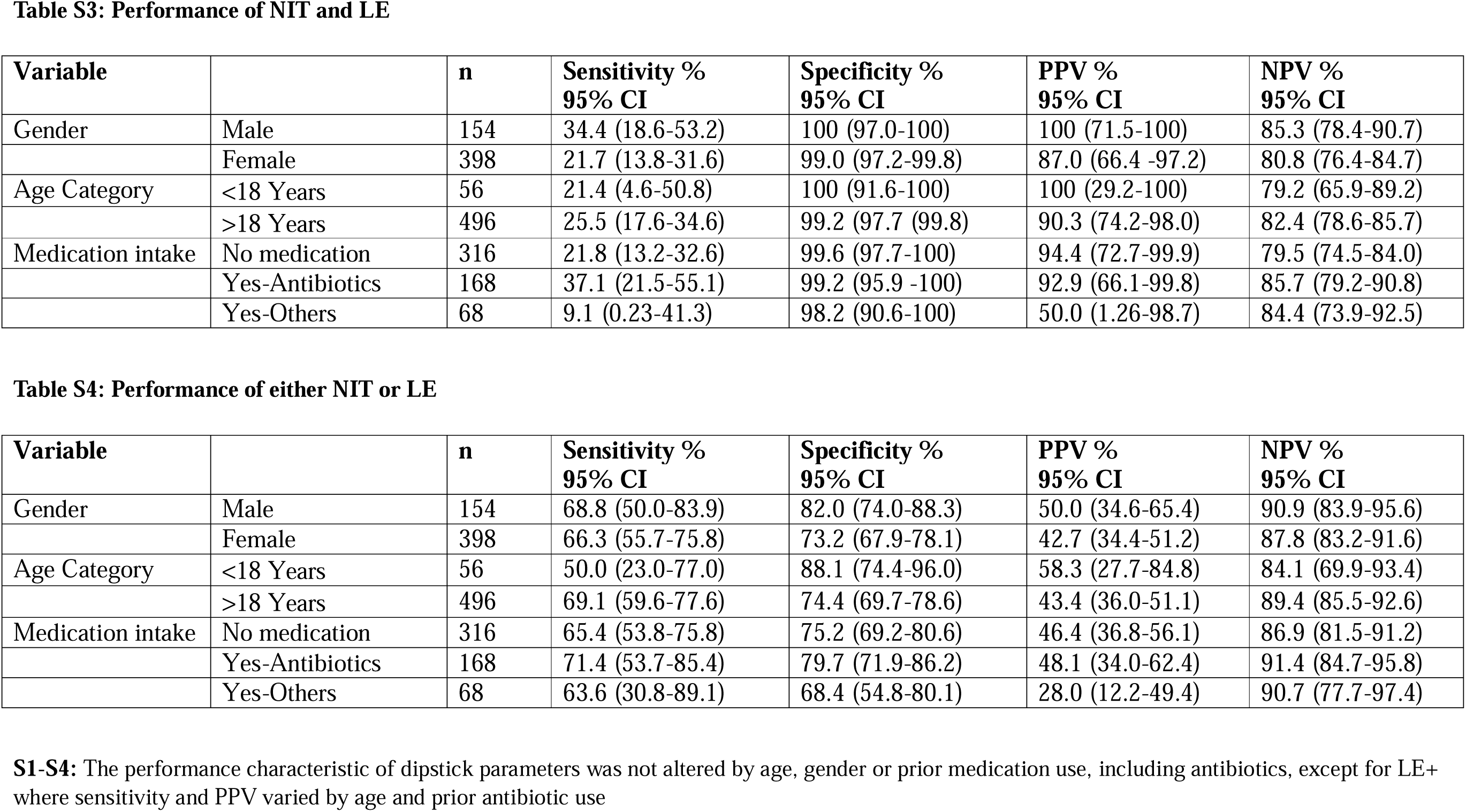
Performance of dipstick parameters across study variables.

## Notes

### Competing Interest Statement

The authors have declared no competing interest.

### Funding Statement

This study was funded by Global challenges research fund (GCRF)

### Author Declarations

University of St. Andrews Teaching and Research Ethics Committee (UTREC), [Approval code. MD15749]; Jomo Kenyatta University of Agriculture and Technology Institutional Ethics Review Board (JKUAT-IERB) [Approval no. JKU/IERC/02316/0166]; National Commission for Science Technology and Innovation (NACOSTI) [Approval no. P/21/12520].

### Summary of Updates

This manuscript has been updated by inccoporating the patient clinical symptoms which was previously missing.An analyis of symptoms and dipstick urinalysis, both individually and in combination have been included.

